# From Sequences to Strategies: Early Detection of New SARS-CoV-2 Variants via Genetic Distance to Reduce Hospitalizations

**DOI:** 10.1101/2025.09.03.25334908

**Authors:** Marika D’Avanzo, Aung Pone Myint, Giacomo Cacciapaglia, Stefan Hohenegger, Francesco Conventi, Marta Nunes

**Author notes:** These authors contributed equally to this article and share first authorship (Marika D’Avanzo, Aung Pone Myint).

## Abstract

The COVID-19 pandemic highlighted the critical need for robust methods to monitor viral evolution and detect emerging variants of concern (VOCs). Traditional genomic surveillance often lacks predictive power. This study expanded an unsupervised machine learning clustering algorithm, based on SARS-CoV-2 Spike protein Levenshtein distance, to track and predict variant predominance across six European countries from 2020 to January 2024. We also investigated the influence of genetic distances and containment strategies on hospitalization rates. Sequences were transformed into temporal chains, and growth parameters were extracted via sigmoid fitting. A deep neural network (DNN) was trained to classify emerging chains as likely dominant, while a CatBoost model assessed variable importance for predicting weekly hospitalizations in Denmark. Simulations explored modifying vaccine genetic distance, containment measures, and VCR.

Approximately 5,000 sequences per week enabled early chain detection within four weeks. The DNN achieved near-perfect classification of chain predominance within 3-4 weeks of appearance. Genetic distances within consecutive chains and with vaccine strains were significant predictors of hospitalizations. Simulations suggest that better-matched vaccines or stricter containment measures could reduce hospitalizations. Doubling vaccination coverage alone had minimal effect but showed additional reductions when combined with strict containment. This integrated framework demonstrates the utility of combining unsupervised and supervised machine learning for real-time tracking and prediction of SARS-CoV-2 variant dynamics and their impact on public health. Our findings underscore the critical role of genetic distances and effective public health interventions in mitigating the burden of emerging variants, supporting timely genomic surveillance and adaptive public health strategies.

## Introduction

Severe acute respiratory syndrome coronavirus 2 (SARS-CoV-2) has evolved continuously producing variants with different levels of transmissibility, immune escape, and disease severity. Its spike protein is central to host cell entry and continuous accumulation of mutations in the spike protein have led to the emergence of phylogenetically distinct variants[1] which can evade immune responses from prior infections or vaccination[2]. Four of these variants, Alpha, Beta, Delta, and Omicron, have been classified as variants of concern (VOCs) by the World Health Organization (WHO), while others have been designated as variants of interest (VOIs) or variants under monitoring (VUMs)[3]. Since December 2021, Omicron and its sub-lineages have globally dominated the epidemiological dynamics. On May 5, 2023, the WHO declared the end of the COVID-19 public health emergency of international concern while acknowledging the ongoing risks posed by future SARS-CoV-2 evolution[4]. This highlights the importance of continuous viral genomic surveillance to promptly detect and assess new variants that could impact public health, particularly in the context of diverse immunity profiles resulting from varied exposure due to irregular vaccines coverage and prior infections.

To address this need, de Hoffer et al.[5] developed an unsupervised machine learning (ML) algorithm to define new variants by clustering the amino acid sequences of the spike protein of SARS-CoV-2 based on the Levenshtein distance, and a time-binned hierarchical clustering with Ward’s method. Clusters across consecutive time bins are then linked as chains when they contain the same dominant spike sequence. These chains were empirically found to correspond to persistent and potentially epidemiologically significant variants. The approach effectively predicted the Alpha and Delta variants in the United Kingdom.

Levi et al.[6] analyzed data from 30 countries to identify variants associated with over 1,000 cases per million population within a three-month period using Jaccard distance. However, the method’s reliance on retrospective metrics, the maximum weekly case count observed over the full variant duration, limits its applicability for real-time prediction. Nicora et al.[7] used k-mer counts and a one-class Support Vector Machine (SVM) to flag anomalous sequences, though it suffered from a high false-positive rate. Rancati et al.[8] also employed spike k-mer representations with an autoencoder to predict lineages that would exceed 10% of total sequences, but its performance was inconsistent in countries with relatively lower sequencing volumes, such as France and Denmark. More recently, Feng et al.[9] developed a transformer-based model designed to forecast future lineage frequencies up to two months in advance, however, it relies on prior lineage designation and focuses exclusively on frequency forecasting.

While COVID-19 vaccines have been shown to protect against infections and hospitalizations[10], the Omicron variant with extensive spike mutations has caused substantial hospitalizations[11] despite lower intrinsic virulence[12]. Current approaches for assessing vaccine effectiveness against novel variants (primarily in vitro neutralization assays[13,14]) have limitations in their timeliness and predictive capacity. Studies such as Cao et al.[15] have explored the relationship between genetic divergence and vaccine effectiveness, suggesting the genetic distance between circulating and vaccine strains as an indicator for immune escape potential. However, a direct, quantitative link between genetic divergence and real-world clinical outcomes has not been established.

In this study, we extend the unsupervised ML algorithm by de Hoffer et al.[5] to analyze spike protein sequences from six European countries. We assess the robustness of the algorithm across geographically and temporally diverse datasets, identify the optimal sequence volume required for early variant detection and evaluate predictive parameters for variant predominance. Furthermore, we incorporate a deep learning classifier to improve early predictions of variant dominance based on early prevalence trends. Additionally, we investigate impact of genetic distance between variants and distances with vaccine strains on observed hospitalizations by leveraging comprehensive Danish health data, together with different containment strategies. By integrating the identification of emerging variants through genetic surveillance with a novel analysis of the direct link between spike protein evolution and hospitalization, this study offers a valuable framework for predicting and responding to future epidemics caused by novel viruses.

## Methods

### Data sources

We analyzed SARS-CoV-2 Spike protein sequences from Germany, Italy, Sweden, Denmark, France, and Spain sourced from the GISAID database[16]. The Oxford Covid-19 Government Response Tracker provided data on Containment and Health Index (CHI) through the end of 2022[17]. Weekly hospitalization numbers, reinfection percentages and weekly vaccination coverage (VCR) in Denmark were obtained from the Denmark Statens Serum Institut (SSI)[18]and complemented by the European Centre for Disease Prevention and Control (ECDC)[19]. Variant-specific hospitalization risks were obtained from a retrospective study in Washington State covering December 2020 to January 2022[20].

### Cross-Country Variant Surveillance

Our analysis began with the algorithm developed by A. de Hoffer et al.[5], to identify viral chains with the data from spike protein sequences collected between January 23, 2020 and January 15, 2024. Chains that were observed at least four weeks and had a peak prevalence exceeding 20% were selected for analysis. Chains with the same or similar dominant sequence that represented the same variant were combined. These chains were associated with the respective variants using spike protein sequences available from NCBI (National Center for Biotechnology Information, National Library of Medicine) and the CoVsurver app on GISAID.

We then fitted the frequencies of selected chains (P_c_) using a mathematical function of the time x to model their growth, with parameters L, *b, and a*, and decline, with L_2_, *b*_*2*_, *and a*_*2*_.

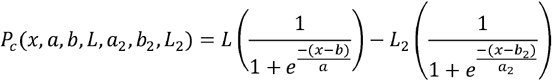

Key parameters derived from this fitting included (*a*) the growth rate, (*b*) the inflection point(point of change in the trend), and (*L*) the upper asymptote (maximum expected prevalence). When fitting only a limited number of weeks *k*, we labelled the parameters with a corresponding subscript (i.e.*a*_*k*_)

We also derived a parameter *t*_0_ that measures the time taken to isolate the chain (*t*_0_ =*b* − 6.91× *a*), and studied its dependence upon the average number of sequences available per week. A schematic representation of the main parameters used in this work has been reported in Supplementary Figure S1).

### Deep Learning-Based Early detection of predominant variant

The dataset was constructed by identifying viral lineages that were observed for at least four consecutive weeks of sequence detection and could be used to derive the parameters needed for the model. Real-world chains from six countries were labeled as the predominant chains if they eventually reached ≥50% prevalence, and the rest as the “transient chains”.

Since real-world data for early-stage variant used to derive the parameters is limited, we applied a data augmentation strategy to generate synthetic chains. For predominant chains, we fitted the key parameters with different distributions from real chains (Supplementary Figure S2), and generated new chains by sampling from these fitted distributions, adding a small noise using an exponential error model. The prevalence at week 1 was computed and the growth curve was recalculated iteratively.

For transient chains, we classified them into two transient chain groups (Supplementary Figure S3): above-threshold (showing some growth tendency) and below-threshold (typically vanishing early). Weekly prevalence differences, expressed as the percentage of chains (*W*_*i* + 1_ − *W*_*i*_), were computed for both groups and modeled using Gaussian distributions. Chains were generated using an iterative procedure sampling weekly difference, ensuring non-negativity and mimicking early extinction events when the values dropped to zero.

We next applied a deep neural network (DNN) model consisting of an input layer with 12 epidemiological features: prevalence at week 1,2, and 3, early-stage fit parameters (*a*_3_, *b*_3_,*L*_3_,*a*_4_,*b*_4_,*L*_4_), and their first-order derivatives ((*a*_4_^′^, *b*_4_^′^,*L*_4_^′^). It was followed by two hidden layers with 64 and 32 neurons respectively, both Rectified Linear Unit (ReLU) activation. The output layer utilized a sigmoid function for binary classification. Model training was carried out using the Adam optimization algorithm and binary cross-entropy as the loss function. The model was trained for 50 epochs using a batch size of 128. The training dataset was randomly split into 80% training and 20% validation.

To evaluate the impact of the initial prevalence of the chains on the classification performance, we generated three different datasets: case 1 - all chains, case 2 – only predominant chains with 15% of prevalence at week 1, and case 3 – only predominant chains with 2% prevalence at week 1. Each dataset was generated to get 10,000 predominant chains and 10,000 transient chains chosen to allow a reasonable training of the models. DNN models were constructed for each dataset using different feature scenarios like using all twelve input features, prevalence values + *a*_3_, *b*_3_,*L*_3_, or prevalence values only.

Model performance was evaluated using the Receiver Operating Characteristic (ROC) curve, the False Positive Rate (FPR) at high True Positive Rate (TPR) thresholds, the confusion matrices, and the signal-to-noise score distribution patterns.

### Analysis of hospitalizations by variants in Denmark

To investigate the relationship between SARS-CoV-2 genetic variation and hospitalization rates, we focused on Denmark due to the availability of comprehensive public data. We selected the chains with >3 week prevalence and ≥50% of prevalence at least at one time point to reduce noise from transient and low-prevalent chains. Chains with the same dominating sequences were combined, even when gaps in their detection weeks led the model to classify them as separate. We calculated the Levenshtein distances between dominating sequences of the consecutive chains (LD1). We computed the distance between dominant sequence and the spike sequence used in the corresponding vaccine being employed at the time (LD_vac). We used the Wuhan strain (EPI_ISL_402124) sequence for the 2020 to 2021-22 season, the BA.5 strain (EPI_ISL_14026118) for 2022-2023 season as both BA.1 and BA.4/5 bivalent vaccines were introduced in that season[21,22], and the XBB.1.5 strain (EPI_ISL_16134259) for the 2023/24 season[23]. To estimate hospitalizations attributable to each variant chain, we assumed that the proportion of sequences belonging to a given chain in any week reflected its proportion of hospitalizations, which were then log-transformed. Other variables included in the analysis were 2-week lagged VCR if primary or booster doses were reported, 2-week lagged CHI which were projected to be 21.43 beyond 2022 as this magnitude had been sustained since April 2022, interaction of the lagged CHI with the detection week (CH_week), and hospitalization risk due to variants in which sub-lineages of Omicron were considered as similar[24–27].

We used a CatBoost regression model to predict weekly hospitalizations per chain. The dataset was randomly split into 80% for training and 20% for testing. We used 5-fold cross-validation and fine-tuned the model hyperparameters with 2000 iterations. Finally, the model’s predictive performance was assessed by comparing predicted hospitalizations to observed values, examining residual plots, and calculating R^2^ and mean absolute error (MAE). Feature importance scores, SHAP (SHapley Additive exPlanations) plot, and partial dependence plots were generated to understand each variable’s contribution.

Finally, we used the trained model to explore the impact of different scenarios: employing better-matched vaccine (distance at 5 for those with ≥15), implementing sustained strong containment measures (maximum observed CHI at 68.57) or temporarily for 8 weeks, doubling VCR, and combinations of high CHI and doubling VCR. All scenarios except the first one were simulated to take actions at week 4 and impact observed at week 6 due to 2-week lag.

We assessed the simulated impact of each scenario by comparing the predicted outcomes with observed hospitalizations. We used 1000 bootstrap simulations to generate 95% confidence intervals (CIs). The results were reported as weekly, cumulative, and percentage reductions.

## Results

### Cross-Country Variant Surveillance

A total of 2862331 sequences for the six countries were analyzed, resulting into 109 chains for Germany, 142 chains for Italy, 95 chains for Sweden, 82 chains for Denmark, 89 chains for France and 98 chains for Spain. A total of 10 chains from Germany, 11 chains from Italy, 8 chains from Sweden, 8 chains from Denmark, 7 chains from France and 12 chains from Spain were selected and successfully matched with the circulating variants.

Comparison of the fit results for the prevalence time evolution for the six considered European countries is reported in Figure 1. The direct comparison of the six main variants across various countries highlighted that the time evolution dynamics of each variant was largely independent of the geographic location (Supplementary Figure S4).

**Figure 1.**
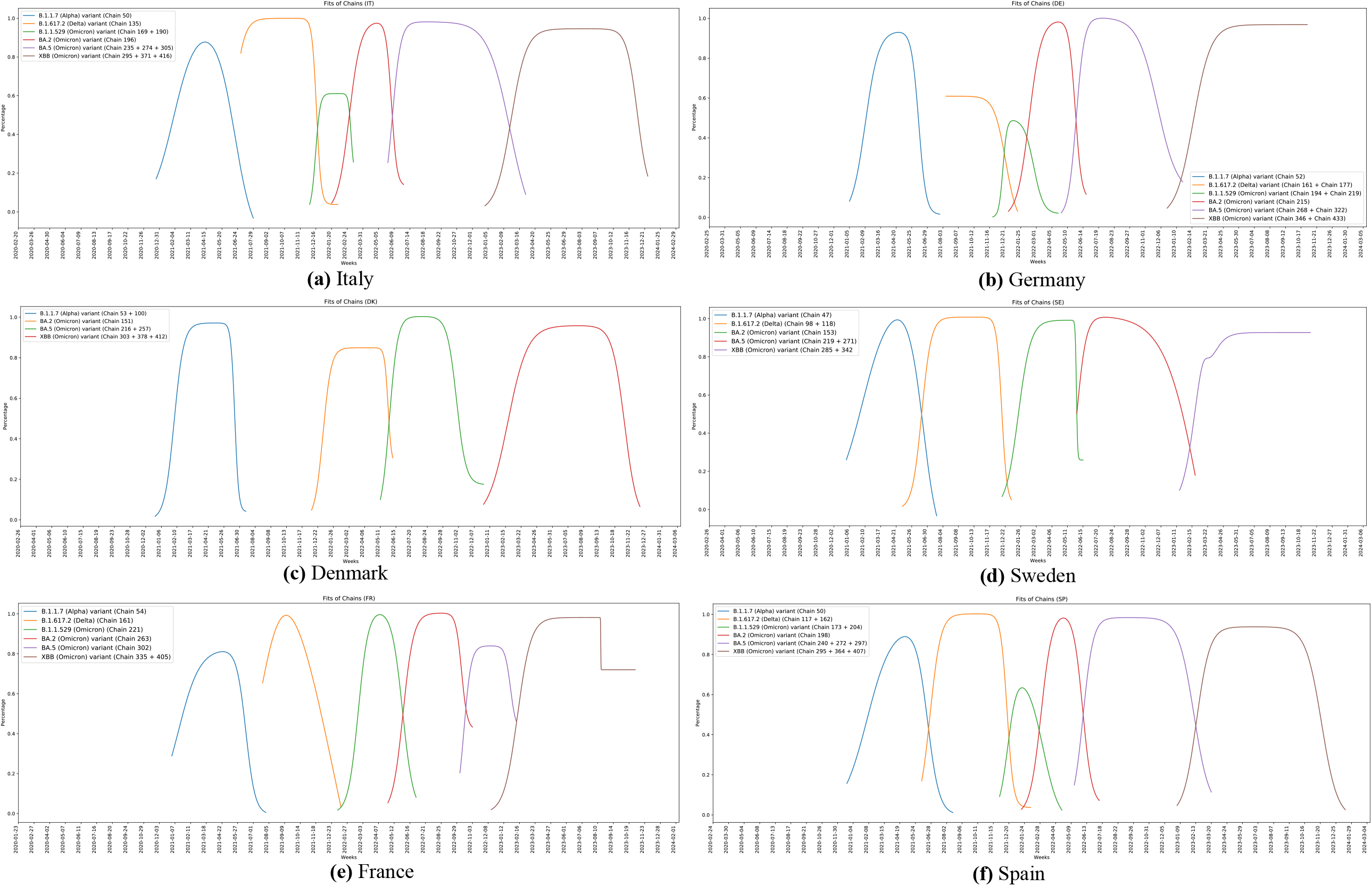
Fitted chains for Italy (a), Germany (b), Denmark (c), Sweden (d), France (e), and Spain (f) from 2020 to 2023. For each country, chains representing the evolution of a variant were fitted using a combination of two sigmoid functions to capture the increase and decrease phase of variant prevalence. On the x-axis, time in weeks is plotted, while on the y-axis the percentage of prevalence (from 0 to 1) of the variant of the chain with respect to all the sequences is considered. Chains are numbered by the algorithm and numbers are assigned according to the number of the first cluster of that chain [5].

The growth rate parameters obtained from the fit at weeks 3 and 4 (*a*_3_ and *a*_4_) differed between dominant and transient chains, and may serve as early indicators of a chain’s long-term behavior. In dominant chains such as France chain 221, the parameters a, b, and L showed stability after a few weeks in comparison with non-dominant chains like Denmark chain 169 with ongoing fluctuations (Supplementary Figure S5).

The fitted curve provided the magnitude of sequencing effort required to minimize *t*_0_ and enhance early variant detection (Figure 2), and approximately 5000 sequences per week were needed for the benchmark, *t*_0_ = 4.

**Figure 2.**
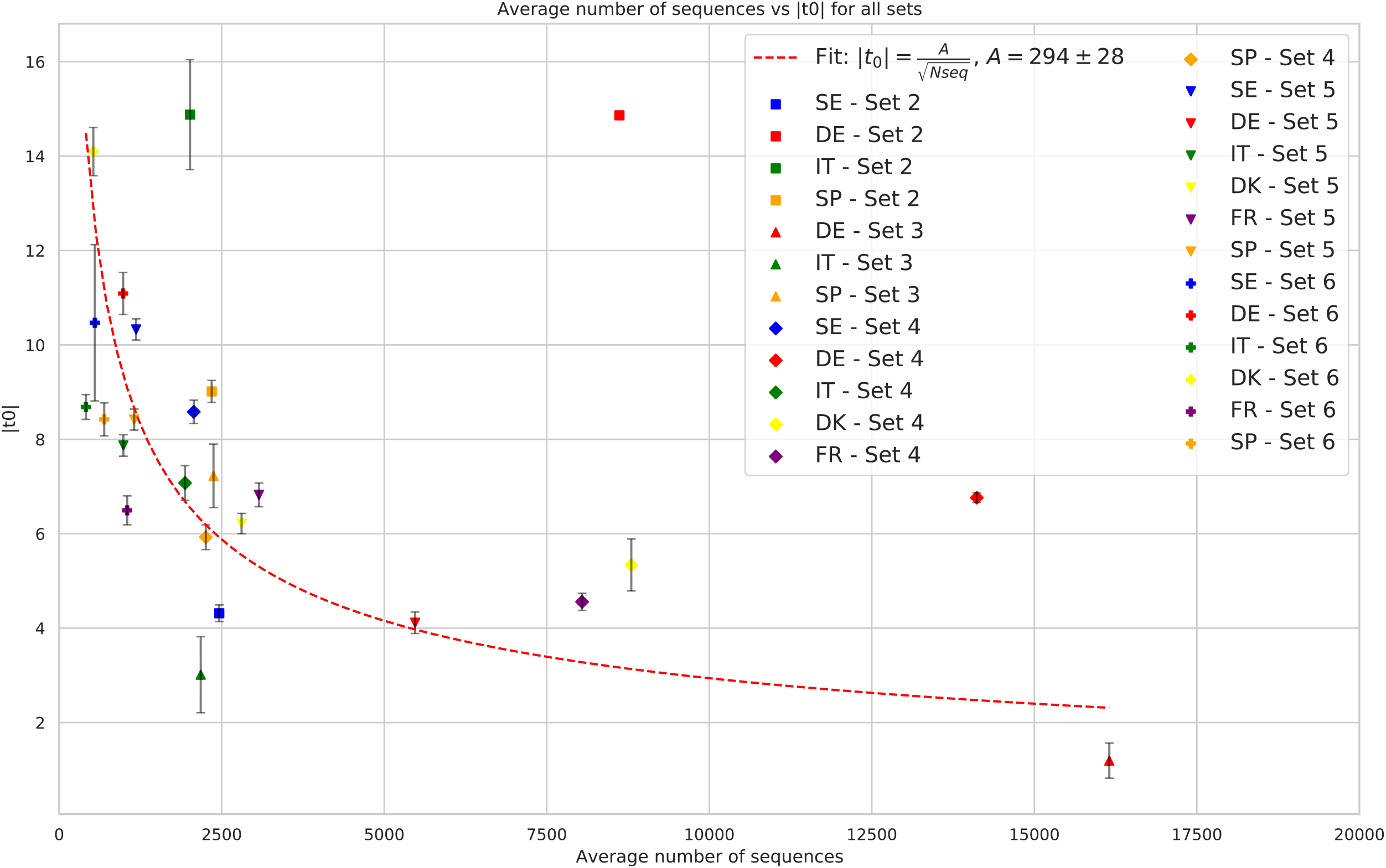
Calibration curve. The average number of sequences available per week is plotted against the absolute value of the first detection time, |*t*_0_|, for variant chains across different country sets. Each point corresponds to a variant detected in a specific country, with marker shape indicating the set and color indicating the country. Error bars represent uncertainty on |*t*_0_| from the fitting procedure. A red dashed curve shows the best fit according to an inverse power law model, 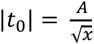, where x is the average weekly number of sequences. The fitted parameter is A = 294 ± 28.

### Deep Learning-Based Early detection of predominant variant

The initial dataset for the DNN model was composed of 37 predominant chains and 78 transient chains. The models trained on the final dataset including simulated chains showed a very low FPR value even at a very high value (99%) of TPR for all the three scenarios using the full set of parameters (Supplementary Table S1). A simplified network obtained adding only growth-related parameters (*a*_3_, *b*_3_,*L*_3_) also significantly improve the prevalence-only model accuracy.

ROC curves confirmed strong overall classification performance, with growth-related parameters offering clear advantages in sensitivity, especially in the more restrictive Case 3 dataset (Supplementary Figure S6). However, relying solely on prevalence values implies a performance drop.

The DNN score distribution further highlighted the effective discriminative power of the deep learning model across all datasets except in the model with prevalence data only (Figure 3). A summary of the classification performance of the deep learning model across all scenarios, considering both different feature subsets and increasing levels of data restriction, can be found in the supplementary Figure S7.

**Figure 3.**
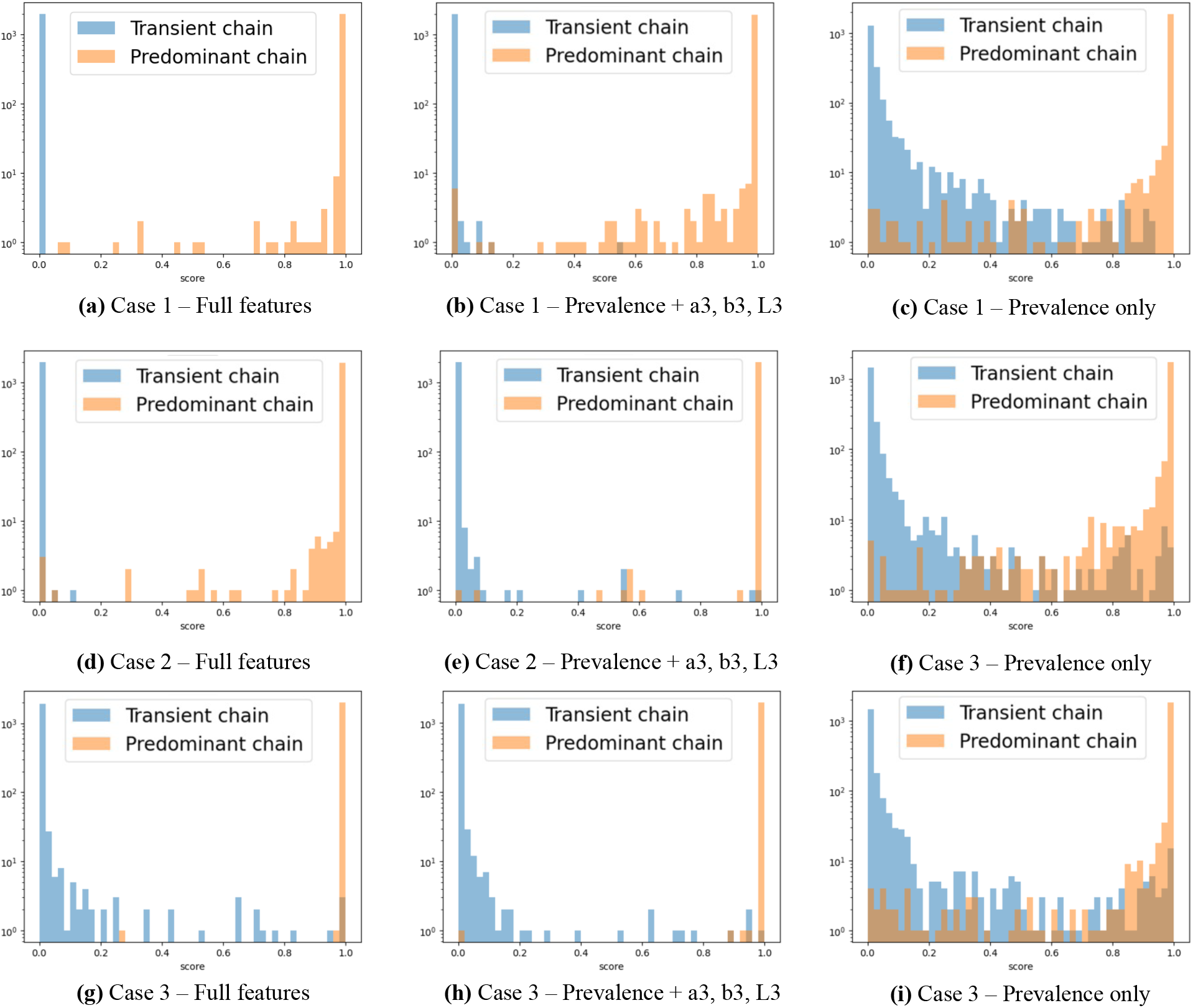
DNN score distribution comparison across dataset configurations and feature sets. The signal-to-noise ratio (i.e. Predominant chains to Transient chains ratio) quantifies the separation between true positive and false positive predictions for different classifier scores. Including growth-related parameters consistently improves the signal-to-noise ratio, particularly for challenging early-stage chains.

### Analysis of hospitalizations by variants in Denmark

14 chains from Denmark were included, and 56,345 (80% of total) hospitalizations were observed by these chains. Weekly hospitalizations by each chain were shown in Figure 4. High distances were observed during the emergence of major variants such as Alpha (chain 53), Delta (105), and the Omicron sublineages BA.2 (151), XBB.1.5 (303), and JN.1 (418) (Figure 4). The reinfection rate, CHI, VCR at each week, and distribution of CH_week can be found in Supplementary Figures S8 and S9.

**Figure 4.**
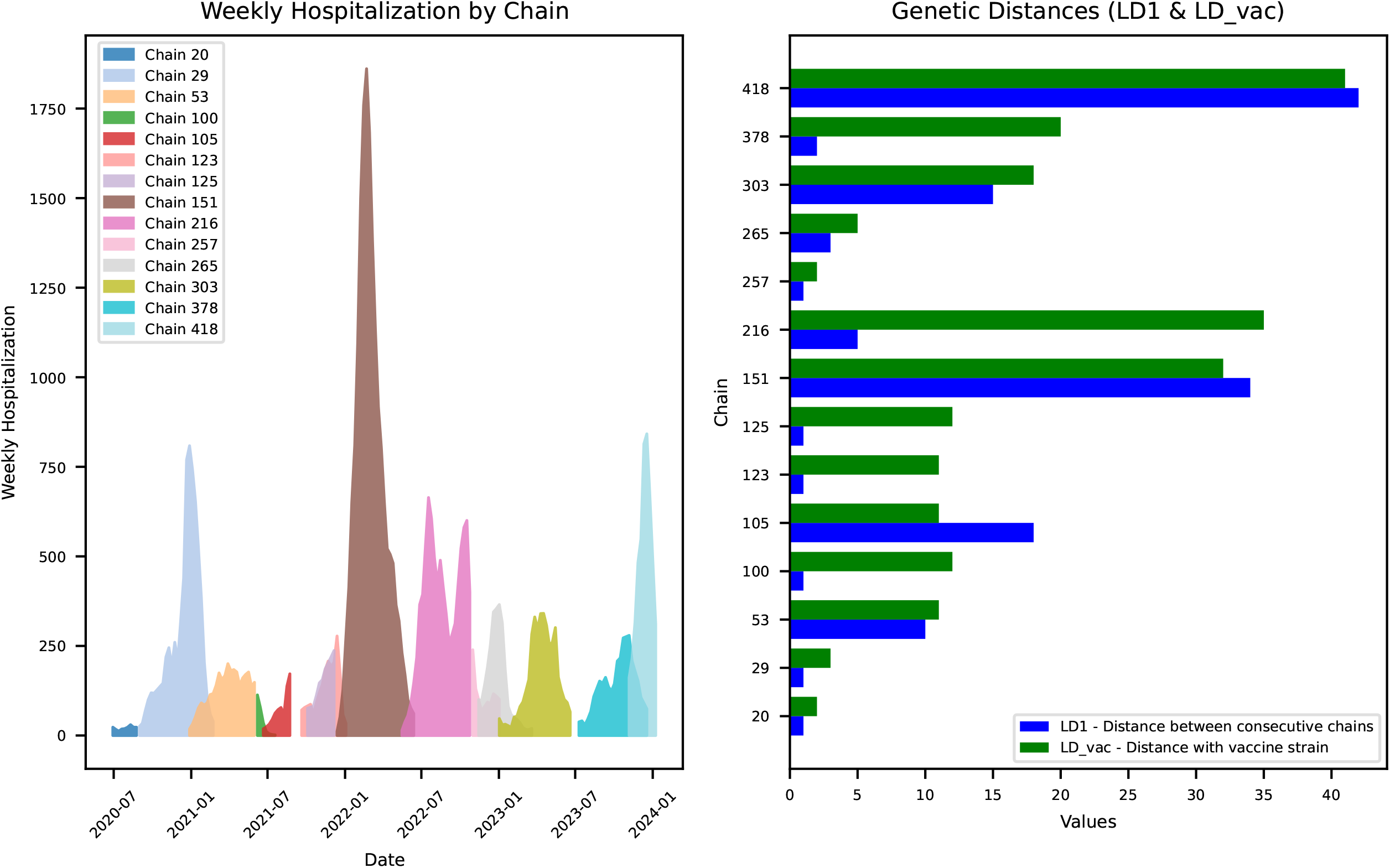
Weekly hospitalizations and Levenshtein distances between consecutive chains and their respective vaccine strain distances for each chain, observed in Denmark from July 2020 to January 2024. Variant classifications: Chain 53 – Alpha, 100 – Delta, 151 – Omicron BA.2, 216 - BA.5, 303 – XBB.1.5, 418 – JN.1.

The CatBoost model demonstrated high predictive accuracy (Supplementary Figure S10), achieving an overall R^2^ score of 0.98 and 0.85 on the unseen test dataset. The MAE was 0.10 for the training set and 0.31 for the test set. Despite the higher error on the test data, the magnitude remained small when compared with the outcome with a minimum of 0.52 and an average of 4.90. Residual diagnostics showed no evidence of heteroscedasticity or non-linearity (Supplementary Figure S10).

The detection week of each chain was the most impactful variable, contributing 26.5% to the model’s explanatory power, followed by LD_vac (15.5%), VCR (14.6%), and LD1 (10.8%) (Figure 5). The SHAP plot (Supplementary Figure S11) indicated that the week counter showed the highest impact on the outcome with top position, and exhibiting bell-shaped association in the partial dependence plot (Supplementary Figure S12). High LD1 and LD_vac values indicated a positive relationship with hospitalizations and high impact on the outcome, while negative association was observed for CHI_lagged. Finally, hospitalizations also showed seasonal trends, increasing during December and January and decreasing during July and August (Supplementary Figure S13).

**Figure 5.**
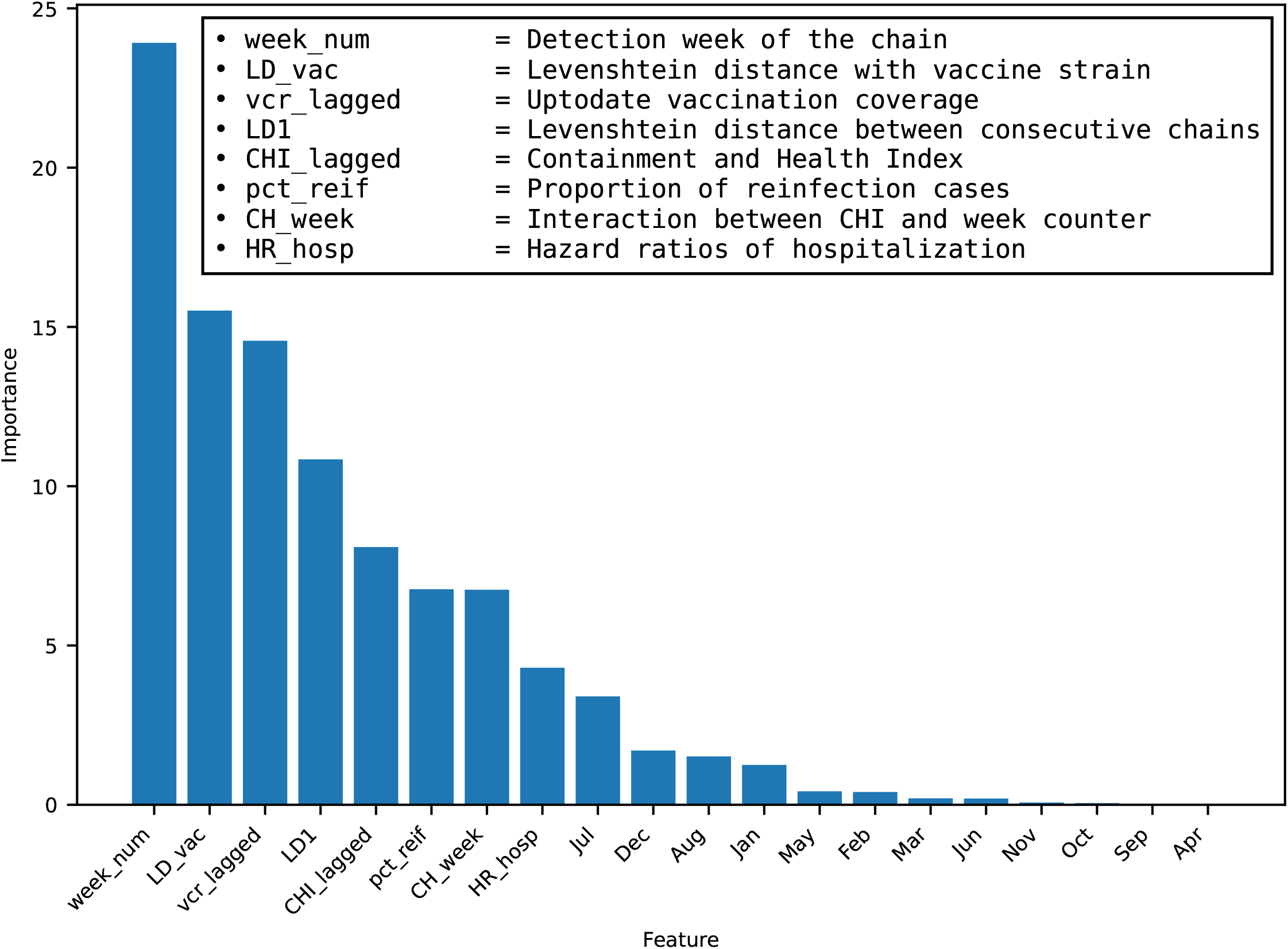
Feature importance of the CatBoost model for prediction of weekly hospitalizations by chains in Denmark (Jul 2020 - Jan 2024).

While simulating with better-matched vaccines, significant reductions in hospitalizations (26-55%) were observed for chains 151, 216, and 418 while nonsignificant reductions observed for chain 303 and no change for chain 378. Temporary or sustained CHI had minimal impact on early waves chain until 123 observed before 2022, since CHI levels were already high during these periods (average CHI >50). Doubling VCR alone produced no statistically significant changes in hospitalization trends across any of the analyzed chains. Additional reductions in hospitalizations were observed in high CHI simulations when combined with doubling VCR.

Details of the simulated impact on the reduction in the percentages from total hospitalizations are presented in Figure 6, and detail simulations of weekly hospitalizations and cumulative hospitalizations can be found in Supplementary Figures S14,S15, and S16.

**Figure 6.**
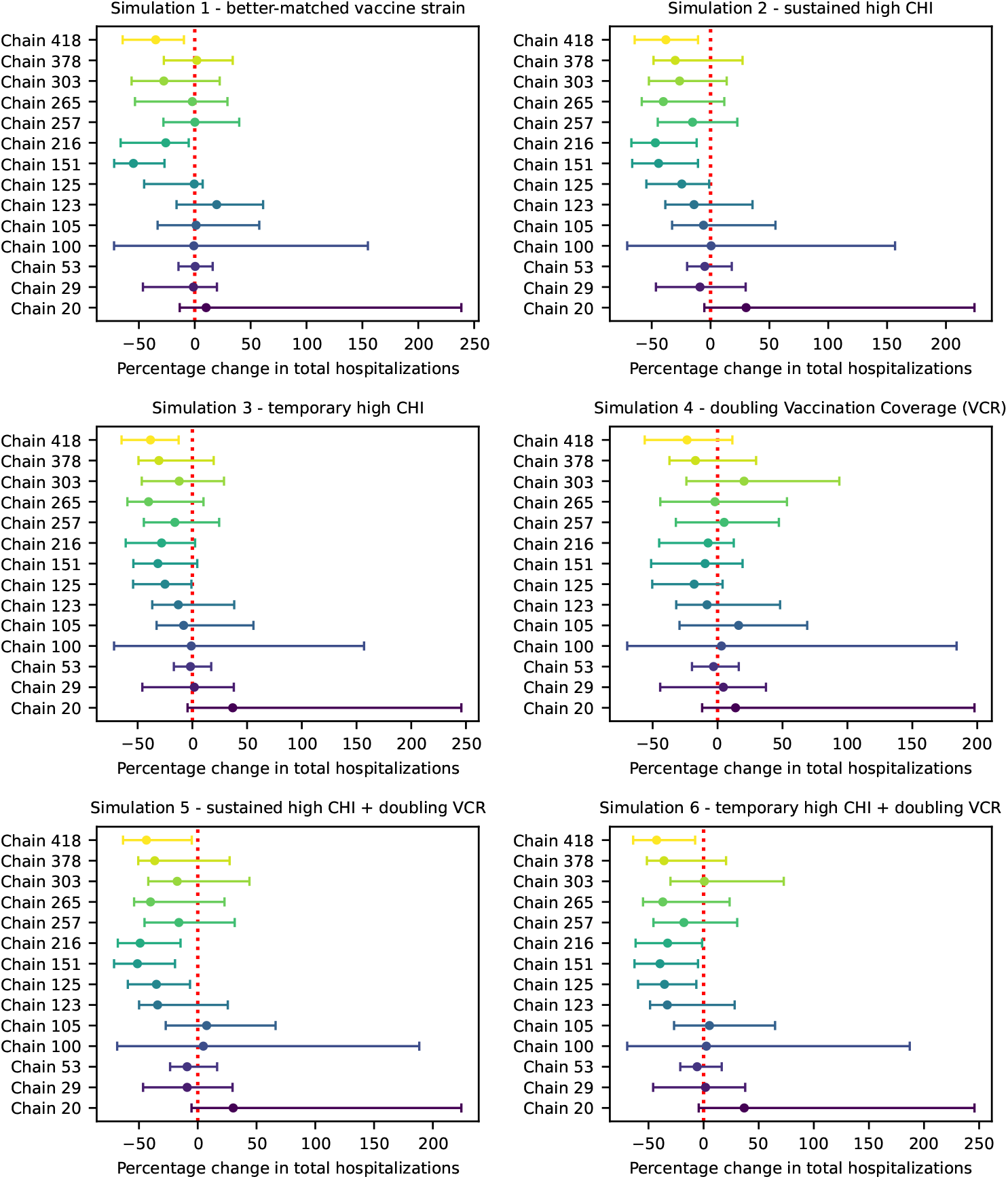
Changes in the proportion of total hospitalizations per chain in Denmark, across different scenarios, with respective 95 % confidence intervals.

## Discussion

This study highlights several critical insights into the optimization of SARS-CoV-2 surveillance strategies and enabling proactive variant detection and response. It was found that approximately 5000 sequences per week for detection within 4 weeks target enables timely detection and facilitates early intervention measures, and provides a universal guideline for sequencing requirements. Currently, reporting of SARS-CoV-2 sequences in GISAID has been diminishing sharply in all countries, and reached lowest point in July 2025 with 7,802 sequences[16].

Unlike traditional surveillance, our DNN model rapidly identified dominant variants using only the first few weeks of prevalence data. It improved this assessment by integrating sigmoid growth parameters and their derivatives. Our findings with low initial-prevalence chains only even showed similar (or even better) classification performance, supporting the idea that early-stage variant detection is feasible even when initial chain prevalence is quite low, which is critical for timely public health responses.

These insights highlight a key advantage of deep learning over rule-based classification or threshold-based methods. Instead of relying on arbitrary cutoffs for identifying concerning variants, our model learned from observed epidemiological dynamics, allowing for data-driven decision-making. Compared to previous studies our model demonstrates improved performance[7,8], usability in real-time monitoring[6], and the adaptability and reliability in scenarios with limited sequencing data[8].

Our study also reinforces the importance of vaccine matching and genetic monitoring in mitigating variant impact. The distances between consecutive variants and distance from the vaccine strain were among the most significant predictors of hospitalization rates, which was supported by reduced vaccine effectiveness for new variants[28,29].

Our simulations indicated a clear link between genetic distance and clinical outcomes at a population level, except in one case, chain 378, which was first reported just before the introduction of the matched XBB.1.5 vaccine. Building upon this, our finding highlights that tracking genetic distance alone can provide actionable insights for hospital capacity planning, even without complete epidemiological data.

Strict non-pharmaceutical interventions showed a significant impact in reduction of hospitalizations, and combining with doubling VCR showed additive benefits, which suggests that a multifaceted approach could yield even greater benefits in the absence of matched vaccines. The observed reductions, particularly in chains with lower initial containment levels or delayed vaccine strain matching, emphasize the importance of timely public health measures in the emergence of genetically distinct variants like chains 151 (BA.2), 216 (BA.5), and 418 (JN.1).

These insights provide a compelling case for sustained investment in genomic surveillance, vaccine development, and integrated public health responses to safeguard against the evolving threat of SARS-CoV-2 and other emerging pathogens.

Several limitations should be taken into account in our study. First, the performance of both the clustering algorithm and the deep learning model was inherently dependent on the availability of genomic data, and may introduce biases due to sequencing delays, particularly during the early emergence of new variants. Second, the predictive power of the DNN relied on features extracted during the first few weeks after a variant’s initial detection from the clustering algorithm, and variants with delayed or irregular dynamics may challenge the model’s classification ability.

Thirdly, the data augmentation strategy might not fully capture the diversity of real-world evolutionary behaviors, particularly for variants exhibiting novel or outlier dynamics. Fourthly, its ability to predict the trajectory of newly emerging variants, especially different viruses, in real-time remains to be systematically validated. Future research should explore integrating real-time data streams, improving robustness against sequencing gaps, and evaluating transferability to other viruses. Another limitation is that the proportion of variants among hospitalized patients might be different from that in the general infected population. However, focusing on major variants in the model may partially address this issue, as these variants are more likely to be associated with higher hospitalization rates. Additionally, we were unable to incorporate detailed population structures of vaccine recipients or hospitalized patients due to data availability constraints. Furthermore, the long-term impact of vaccinations from previous seasons on hospitalization rates was not included in our analysis. While this omission may not significantly alter outcomes, it remains a factor worth exploring in future studies to ensure a more comprehensive understanding. Finally, modifying key variables within the CatBoost model introduced wide CIs resulting many scenarios’ reduction insignificant. Future research should aim to precisely quantify the relationship between the magnitude of variable modification and the resulting predictive accuracy, thereby refining the ability to generate nuanced and robust hospital burden forecasts. This would not only improve the interpretation of scenario-based predictions but also support the development of more resilient and informative forecasting methodologies.

## Conclusion

The integration of optimized sequencing rates, early-stage classification with deep learning, and genetic-based hospitalization risk assessment offers a powerful, data-driven framework for SARS-CoV-2 surveillance. We show that adequate sequencing volume (impacting on the time needed to detect chains) together with classification models based on growth dynamics and leveraging genetic distance as a key predictor can enhance the ability of public health systems to promptly identify and mitigate the impact of new variants. We demonstrate that early variant classification is feasible using only a few weeks of prevalence data and validating the importance of sigmoid-based features in improving classification accuracy even with low initial-prevalence chains. We prove evidence that genetic distance predicts hospitalization risk, supporting real-time response strategies including vaccine adaptation programs and we highlight the need for integrated deep learning, genetic monitoring, and vaccination strategies for pandemic preparedness.

These findings contribute to a growing framework for real-time, adaptive surveillance systems that can rapidly respond to emerging epidemiological threats. Future work will focus on enhancing the robustness of the DNN model, exploring alternative architectures (e.g., transformer-based models), and refining transient chain modeling techniques to further improve predictive accuracy.

## Data Availability

All data produced in the present study are available upon reasonable request to the authors.

## References

1. Jackson CB, Farzan M, Chen B, Choe H. Mechanisms of SARS-CoV-2 entry into cells. Nat Rev Mol Cell Biol. 2022 Jan;23(1):3–20.

2. Markov PV, Ghafari M, Beer M, Lythgoe K, Simmonds P, Stilianakis NI, et al. The evolution of SARS-CoV-2. Nat Rev Microbiol. 2023 Jun;21(6):361–79.

3. WHO. Updated working definitions and primary actions for SARSCoV2 variants [Internet]. 2023 [cited 2025 Jul 23]. Available from: https://www.who.int/publications/m/item/updated-working-definitions-and-primary-actions-for--sars-cov-2-variants

4. WHO. Statement on the fifteenth meeting of the IHR (2005) Emergency Committee on the COVID-19 pandemic [Internet]. 2023 [cited 2024 Aug 26]. Available from: https://www.who.int/news/item/05-05-2023-statement-on-the-fifteenth-meeting-of-the-international-health-regulations-(2005)-emergency-committee-regarding-the-coronavirus-disease-(covid-19)-pandemic

5. de Hoffer A, Vatani S, Cot C, Cacciapaglia G, Chiusano ML, Cimarelli A, et al. Variant-driven early warning via unsupervised machine learning analysis of spike protein mutations for COVID-19. Sci Rep. 2022 Jun 3;12(1):9275.

6. Levi R, Zerhouni EG, Altuvia S. Predicting the spread of SARS-CoV-2 variants: An artificial intelligence enabled early detection. PNAS Nexus. 2024 Jan 1;3(1):pgad424.

7. Nicora G, Salemi M, Marini S, Bellazzi R. Predicting emerging SARS-CoV-2 variants of concern through a One Class dynamic anomaly detection algorithm. BMJ Health Care Inform. 2022 Dec 9;29(1):e100643.

8. Rancati S, Nicora G, Prosperi M, Bellazzi R, Salemi M, Marini S. Forecasting dominance of SARS-CoV-2 lineages by anomaly detection using deep AutoEncoders. Briefings in Bioinformatics. 2024 Nov 1;25(6):bbae535.

9. Feng Y, Goldberg EE, Kupperman M, Zhang X, Lin Y, Ke R. CovTransformer: A transformer model for SARS-CoV-2 lineage frequency forecasting. Virus Evolution. 2024 Oct 17;10(1):veae086.

10. Graña C, Ghosn L, Evrenoglou T, Jarde A, Minozzi S, Bergman H, et al. Efficacy and safety of COVIDLJ19 vaccines - Graña, C - 2022 | Cochrane Library. [cited 2025 Jul 23]; Available from: https://www.cochranelibrary.com/cdsr/doi/10.1002/14651858.CD015477/full

11. WHO. WHO COVID-19 dashboard [Internet]. datadot. [cited 2025 Apr 18]. Available from: https://data.who.int/dashboards/covid19/hospitalizations

12. Carabelli AM, Peacock TP, Thorne LG, Harvey WT, Hughes J, de Silva TI, et al. SARS-CoV-2 variant biology: immune escape, transmission and fitness. Nat Rev Microbiol. 2023 Mar;21(3):162–77.

13. Cromer D, Steain M, Reynaldi A, Schlub TE, Khan SR, Sasson SC, et al. Predicting vaccine effectiveness against severe COVID-19 over time and against variants: a meta-analysis. Nat Commun. 2023 Mar 24;14(1):1633.

14. Khoury DS, Docken SS, Subbarao K, Kent SJ, Davenport MP, Cromer D. Predicting the efficacy of variant-modified COVID-19 vaccine boosters. Nat Med. 2023 Mar;29(3):574–8.

15. Cao L, Lou J, Chan SY, Zheng H, Liu C, Zhao S, et al. Rapid evaluation of COVID-19 vaccine effectiveness against symptomatic infection with SARS-CoV-2 variants by analysis of genetic distance. Nat Med. 2022 Aug;28(8):1715–22.

16. Khare S, Gurry C, Freitas L, Schultz MB, Bach G, Diallo A, et al. GISAID’s Role in Pandemic Response. CCDCW. 2021 Dec 3;3(49):1049–51.

17. Hale T, Angrist N, Goldszmidt R, Kira B, Petherick A, Phillips T, et al. A global panel database of pandemic policies (Oxford COVID-19 Government Response Tracker). Nat Hum Behav. 2021 Apr;5(4):529–38.

18. SSI. SSI’s interaktive dashboards [Internet]. Denmark Statens Serum Institut. [cited 2024 Apr 27]. Available from: https://experience.arcgis.com/template/099eb5c9acea4e18b411997815be2f98

19. ECDC. Data on COVID-19 vaccination in the EU/EEA [Internet]. 2020 [cited 2024 Apr 27]. Available from: https://www.ecdc.europa.eu/en/covid-19/data

20. Paredes MI, Lunn SM, Famulare M, Frisbie LA, Painter I, Burstein R, et al. Associations between SARS-CoV-2 variants and risk of COVID-19 hospitalization among confirmed cases in Washington State: a retrospective cohort study. medRxiv. 2022 Feb 16;2021.09.29.21264272.

21. SSI. Vaccination against COVID-19, influenza and pneumococcal disease [Internet]. 2022 [cited 2025 Jun 2]. Available from: https://en.ssi.dk/news/epi-news/2022/no-35---2022

22. SSI. This autumn’s influenza and COVID-19 vaccination programme, 2023/2024 [Internet]. 2023 [cited 2025 Jun 2]. Available from: https://en.ssi.dk/news/epi-news/2023/no-39---2023

23. SSI. Retningslinje for vaccination mod covid-19 og influenza - historisk [Internet]. 2023 [cited 2025 Jul 23]. Available from: http://www.sst.dk/da/udgivelser/2023/Retningslinje-for-vaccination-mod-covid-19-og-influenza

24. Harman K, Nash SG, Webster HH, Groves N, Hardstaff J, Bridgen J, et al. Comparison of the risk of hospitalisation among BA.1 and BA.2 COVID-19 cases treated with sotrovimab in the community in England. Influenza and Other Respiratory Viruses. 2023;17(5):e13150.

25. Aziz NA, Nash SG, Zaidi A, Nyberg T, Groves N, Hope R, et al. Risk of severe outcomes among SARS-CoV-2 Omicron BA.4 and BA.5 cases compared to BA.2 cases in England. Journal of Infection. 2023 Jul 1;87(1):e8–11.

26. WHO. XBB.1.5 Updated Rapid Risk Assessment [Internet]. 2023 Jan. Available from: https://www.who.int/docs/default-source/coronaviruse/25012023xbb.1.pdf?sfvrsn=c3956081_1

27. WHO. JN.1 variant update and risk evaluation [Internet]. 2024 Apr. Available from: https://www.who.int/docs/default-source/coronaviruse/15042024_jn1_ure.pdf?sfvrsn=8bd19a5c_7

28. Gram MA, Emborg HD, Schelde AB, Friis NU, Nielsen KF, Moustsen-Helms IR, et al. Vaccine effectiveness against SARS-CoV-2 infection or COVID-19 hospitalization with the Alpha, Delta, or Omicron SARS-CoV-2 variant: A nationwide Danish cohort study. PLOS Medicine. 2022 Sep 1;19(9):e1003992.

29. Moustsen-Helms IR, Bager P, Larsen TG, Møller FT, Vestergaard LS, Rasmussen M, et al. Relative vaccine protection, disease severity, and symptoms associated with the SARS-CoV-2 omicron subvariant BA.2.86 and descendant JN.1 in Denmark: a nationwide observational study. The Lancet Infectious Diseases. 2024 Sep 1;24(9):964–73.

