## Supplementary for "From Sequences to Strategies: Early Detection of New SARS-CoV-2 Variants via Genetic Distance to Reduce Hospitalizations"

### Supplementary materials

This supplementary material is hosted by *Eurosurveillance* as supporting information alongside the article “From Sequences to Strategies: Early Detection of New SARS-CoV-2 Variants via Genetic Distance to Reduce Hospitalizations”, on behalf of the authors, who remain responsible for the accuracy and appropriateness of the content. The same standards for ethics, copyright, attributions and permissions as for the article apply. Supplements are not edited by *Eurosurveillance* and the journal is not responsible for the maintenance of any links or email addresses provided therein.

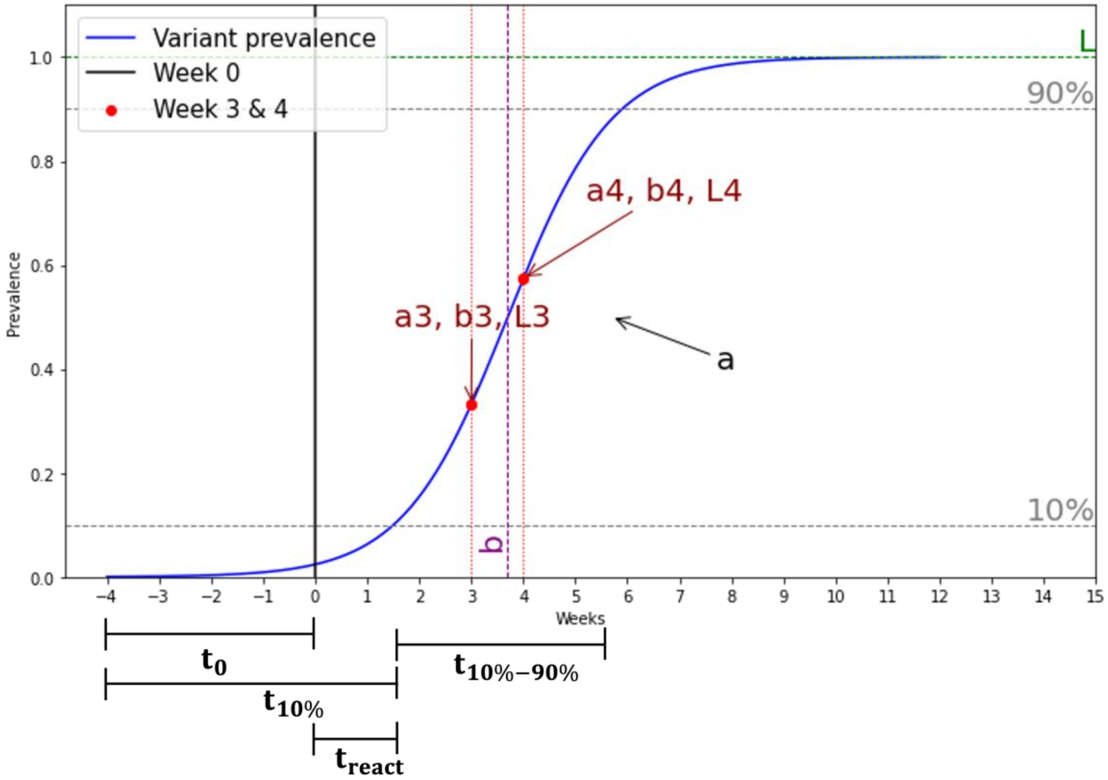


Supplementary Figure S1. Visual representation of the derived parameters used to characterize the growth dynamics of a SARS-CoV-2 variant based on a sigmoid curve. The logistic function models the weekly proportion of genomes attributed to a given variant. L represents the upper asymptote (maximum prevalence). The slope parameter a corresponds to the growth rate, while b indicates the inflection point of the curve. Temporal metrics include: $t_0$ (delay from first occurrence to algorithmic detection), $t_{10\%}$ (time to reach 10% prevalence), $t_{react}$ (reaction time), and $t_{10\%-90\%}$ (time span between 10% and 90% prevalence). Early parameters a, b and L obtained at week 3 and at week 4 are also highlighted in the figure.

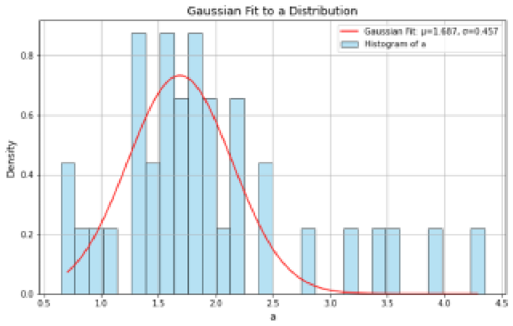


**(a)** Fit to a parameter distribution

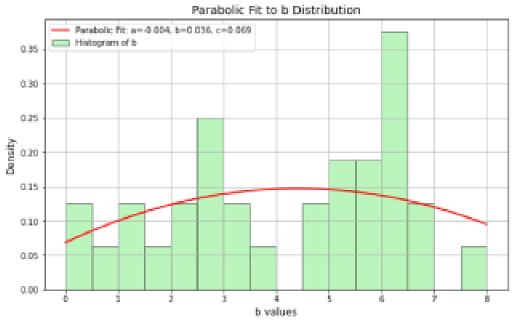


**(b)** Fit to b parameter distribution

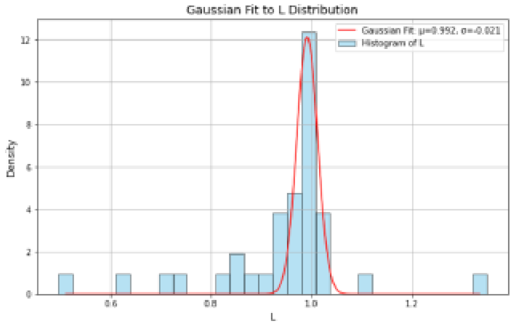


**(c)** Fit to L parameter distribution

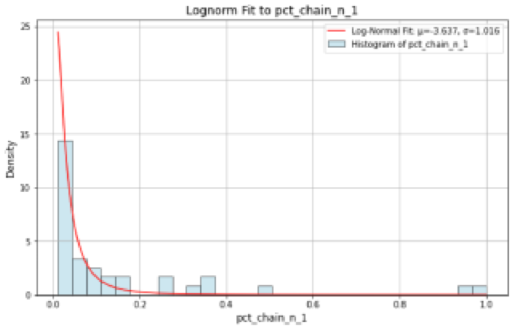


**(d)** Fit to *pct_chain_n_1* parameter distribution

Supplementary Figure S2. Fit procedure applied to key parameters distributions. The parameter a was fitted with a Gaussian distribution (a), the parameter b with a parabolic distribution (b), the parameter L with a Gaussian distribution and the parameter pct_chain_n_1 with a lognorm distribution (d).

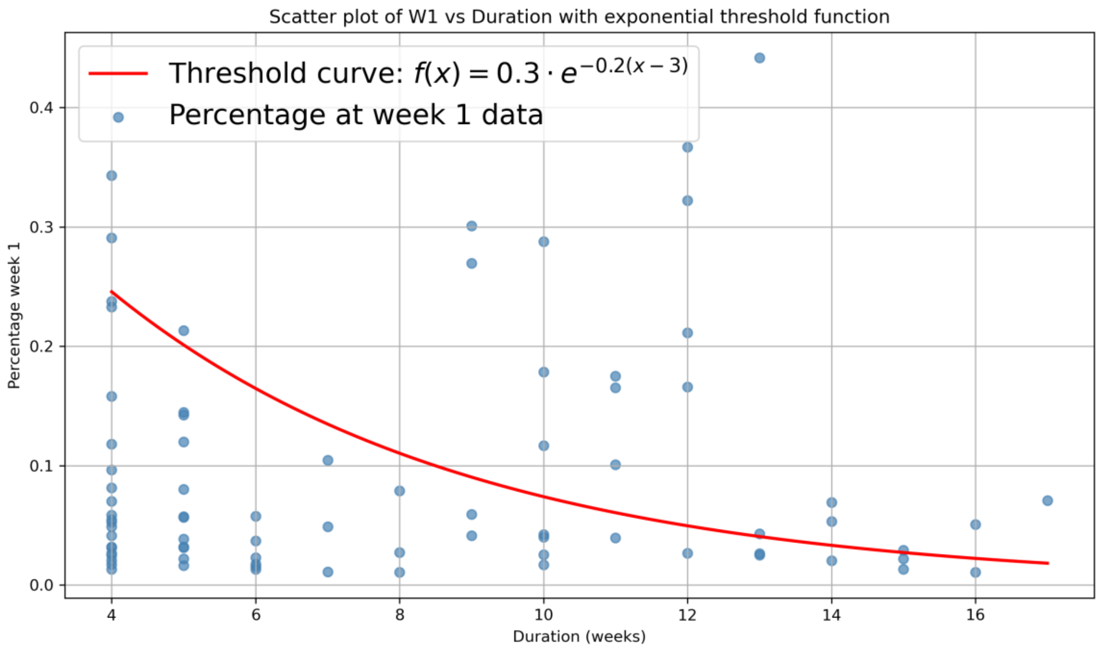


Supplementary Figure S3. Percentage of prevalence at Week 1 vs Duration of real Transient chains. Real transient chains were classified based on their duration and initial prevalence. The prevalence vs duration distribution highlighted two different phase space regions. A cut on this distribution has been applied using an exponential threshold function defining two transient chain groups: above-threshold (showing some growth tendency) and below-threshold (typically vanishing early).

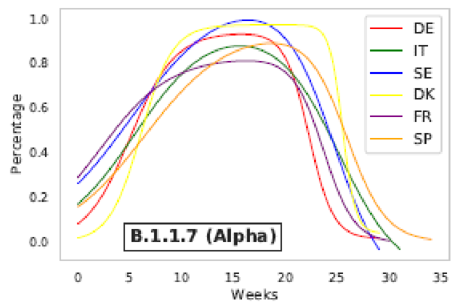


**(a)** Set 1

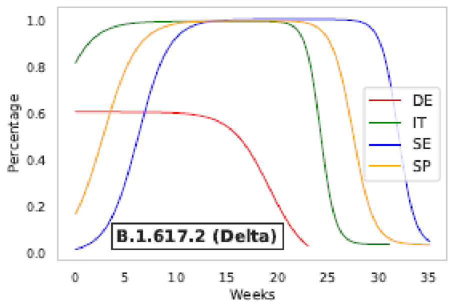


**(b)** Set 2

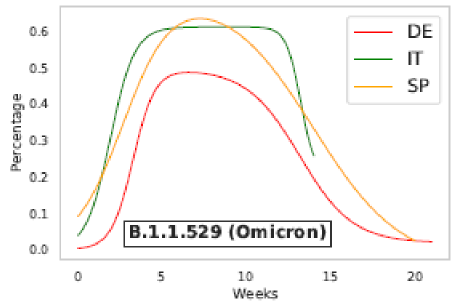


**(c)** Set 3

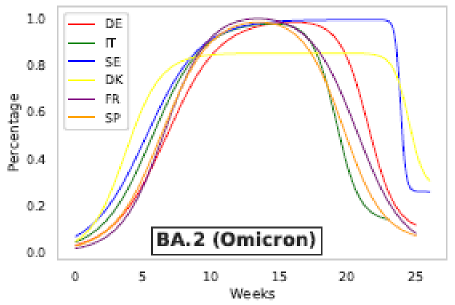


**(d)** *Set 4*

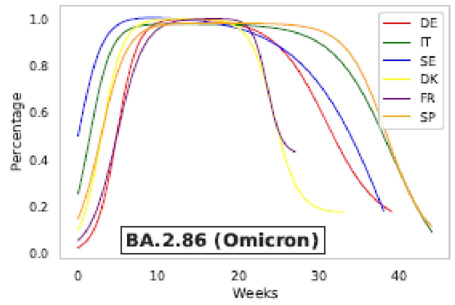


**(e)** Set 5

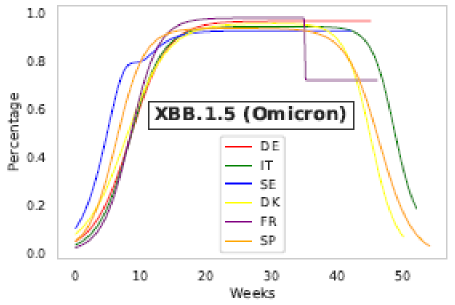


**(f)** Set 6

Supplementary Figure S4: Six sets of fitted chains. (a) Set 1. B.1.1.7 (Alpha) variant. Period: 2020W51 to 2021W33. (b) Set 2. B.1.617.2 (Delta) variant. Period: 2021W19 to 2022W06. (c) Set 3. B.1.1.529 (Omicron) variant. Period: 2021W48 to 2022W17. (d) Set 4. BA.2 (Omicron) variant. Period: 2021W50 to 2022W28. (e) Set 5. BA.5 (Omicron) variant. Period: 2021W50 to 2022W28. (f) Set 6. XBB (Omicron) variant. Period: 2022W52 to 2024W00.

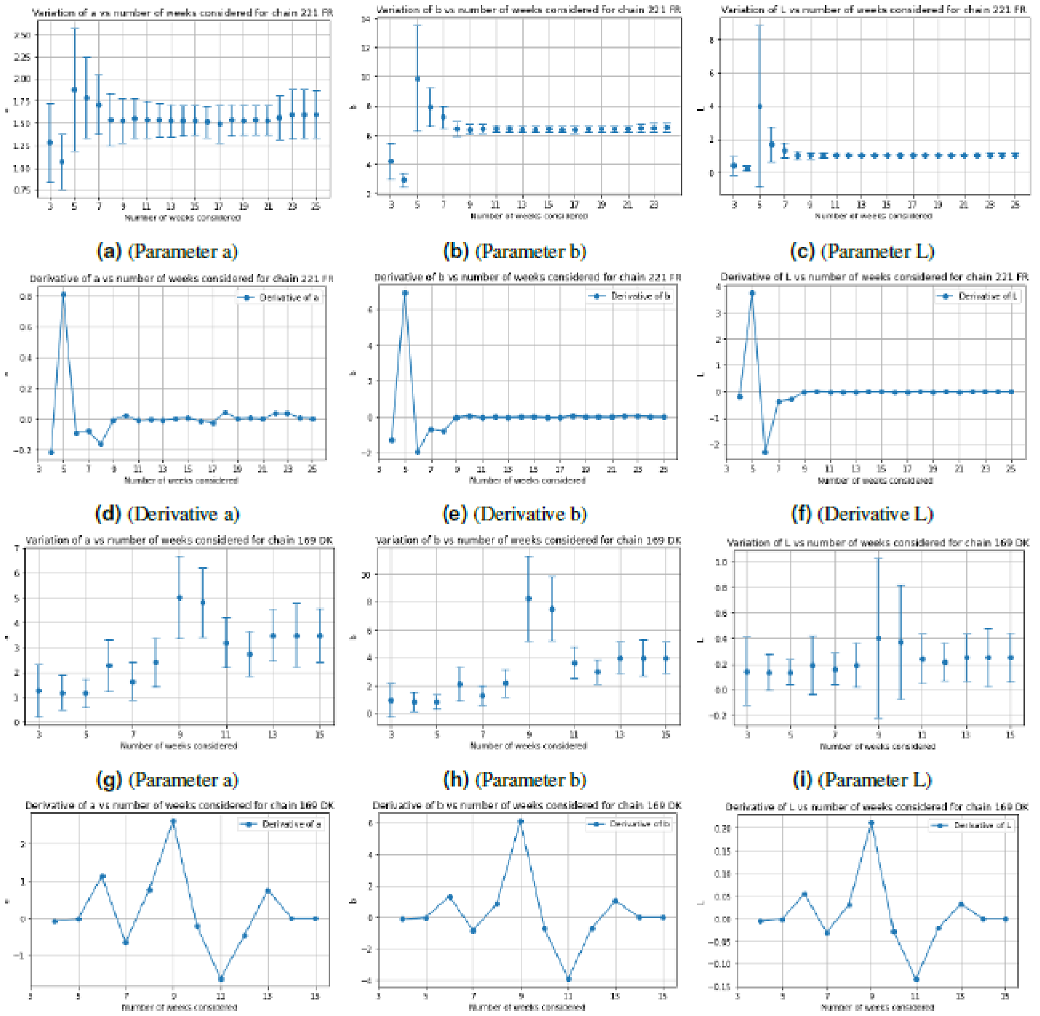


(a) (Parameter a) (b) (Parameter b) (c) (Parameter L)

(d) (Derivative a) (e) (Derivative b) (f) (Derivative L)

(g) (Parameter a) (h) (Parameter b) (i) (Parameter L)

(j) (Derivative a) (k) (Derivative b) (l) (Derivative L)

Supplementary Figure S5. Evolution of the a, b, and L parameters and their derivatives over increasing time windows. Top two rows (a-f) refer to a dominant chain (France, chain 221), and bottom two (g-l) to a non-dominant chain (Denmark, chain 169).

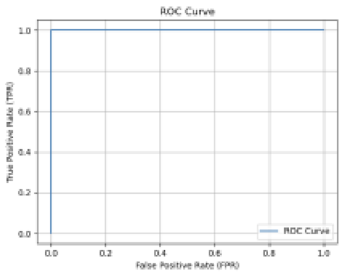


(a) Case 1 – Full features

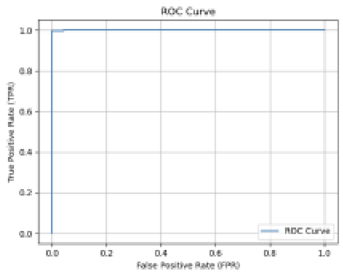


(b) Case 1 – Prevalence + a3, b3, L3

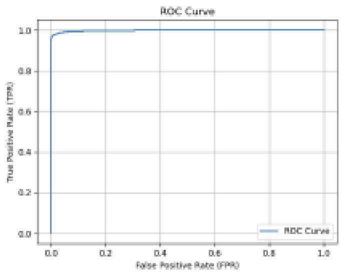


(c) Case 1 – Prevalence only

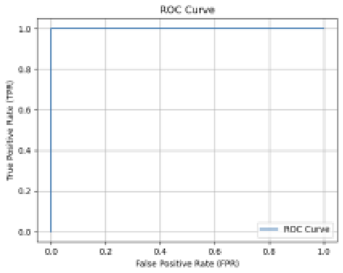


(d) Case 2 – Full features

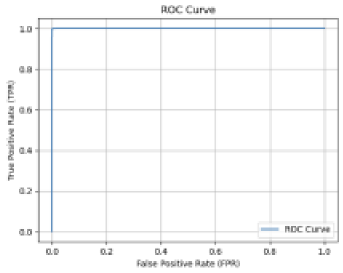


(e) Case 2 – Prevalence + a3, b3, L3

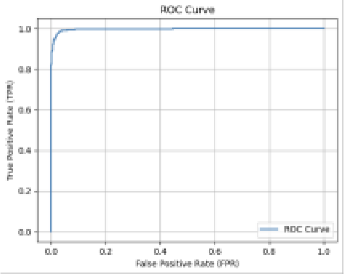


(f) Case 2 – Prevalence only

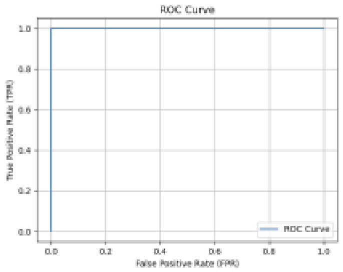


(g) Case 3 – Full features

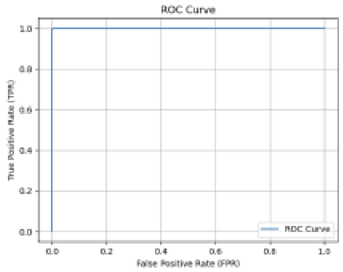


(h) Case 3 – Prevalence + a3, b3, L3

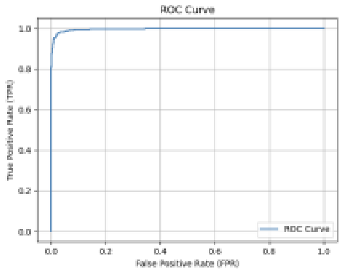


(i) Case 3 – Prevalence only

Supplementary Figure S6. Receiver Operating Characteristic (ROC) curves for the deep learning model under different dataset restrictions and feature sets. Rows represent dataset scenarios: Case 1 (no restriction), Case 2 (pct_chain_n_1<0.17), and Case 3 (pct_chain_n_1<0.02). Columns compare three feature configurations: full feature set (including prevalence and sigmoid-based parameters), prevalence + a_3, b_3, L_3, and prevalence only. The model achieves excellent discrimination performance, especially when growth-related parameters are included.

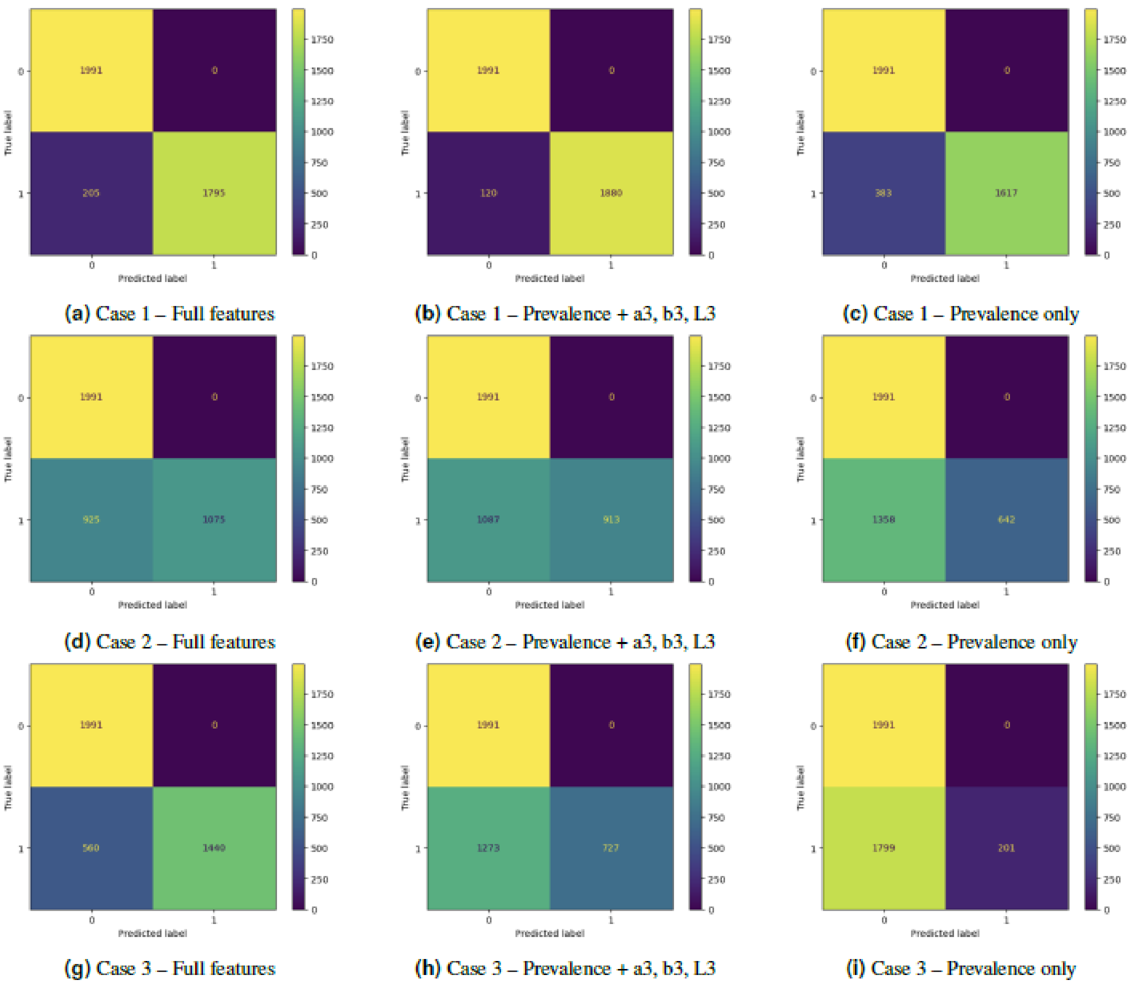


Supplementary Figure S7. Confusion matrices for the deep learning model across all dataset scenarios. Each matrix reports the classification performance under the three levels of data restriction (Cases 1–3) and the three feature sets. Including growth-related parameters leads to a reduction in false positives, especially in the most restrictive scenarios, highlighting the importance of early dynamics for robust prediction

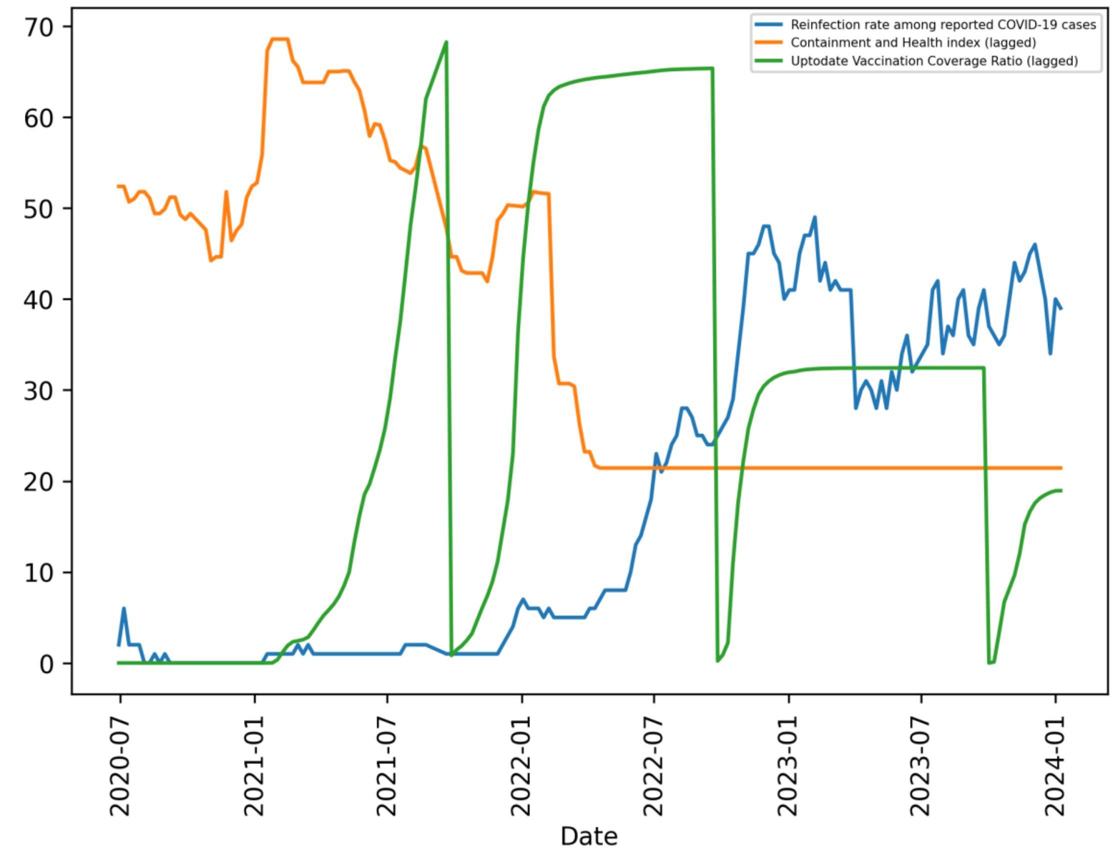


Supplementary Figure S8. Reinfection percentage among reported COVID-19 cases, containment and health index, and up-to-date COVID-19 vaccination coverage of Denmark

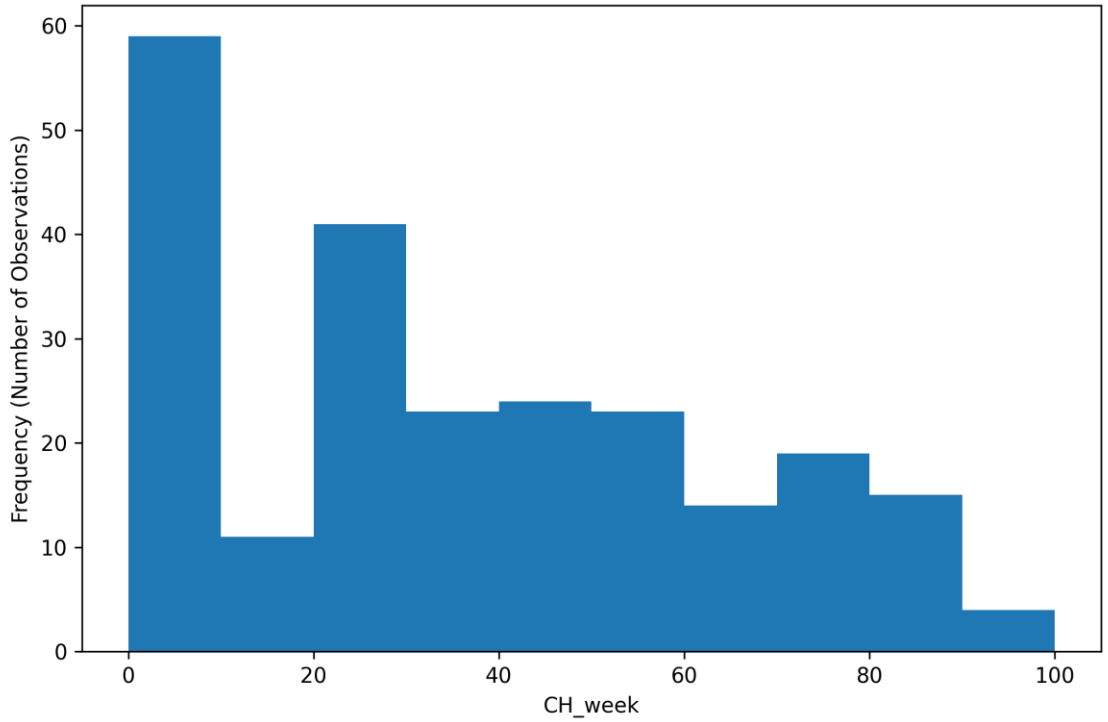


Supplementary Figure S9. Histogram showing the frequency distribution of the variable CH_week, which captures the interaction between containment and health index and the week counter of chains for Denmark from July 2020 to January 2024 (scaled 0 - 100)

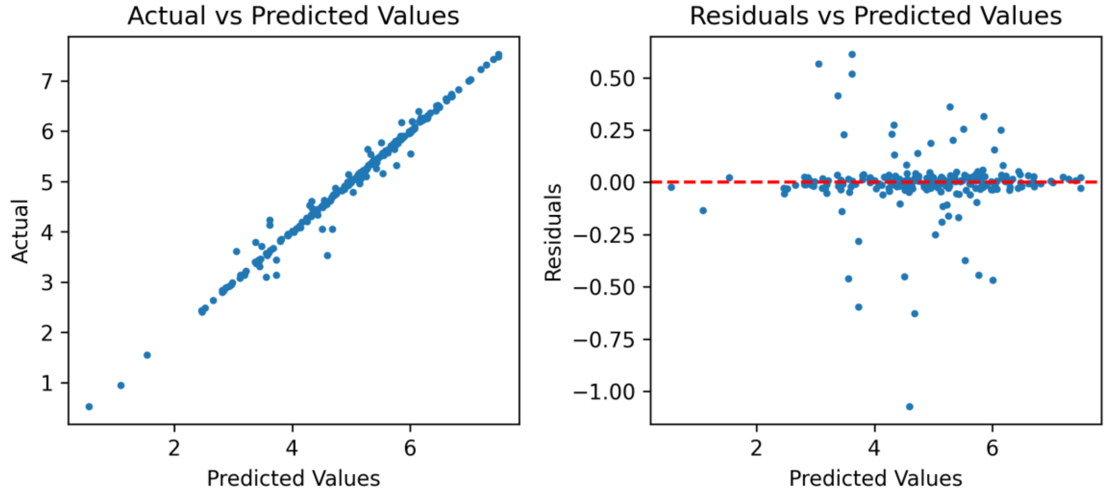


Supplementary Figure S10. Performance Evaluation of CatBoost model for predicting weekly hospitalizations in Denmark. Left panel: Actual vs. predicted values, showing a strong linear pattern indicating high prediction accuracy. Right panel: Residuals vs. predicted values, where the random spread around zero (red dashed line) suggests homoscedasticity, unbiased errors, and a proper model fit.

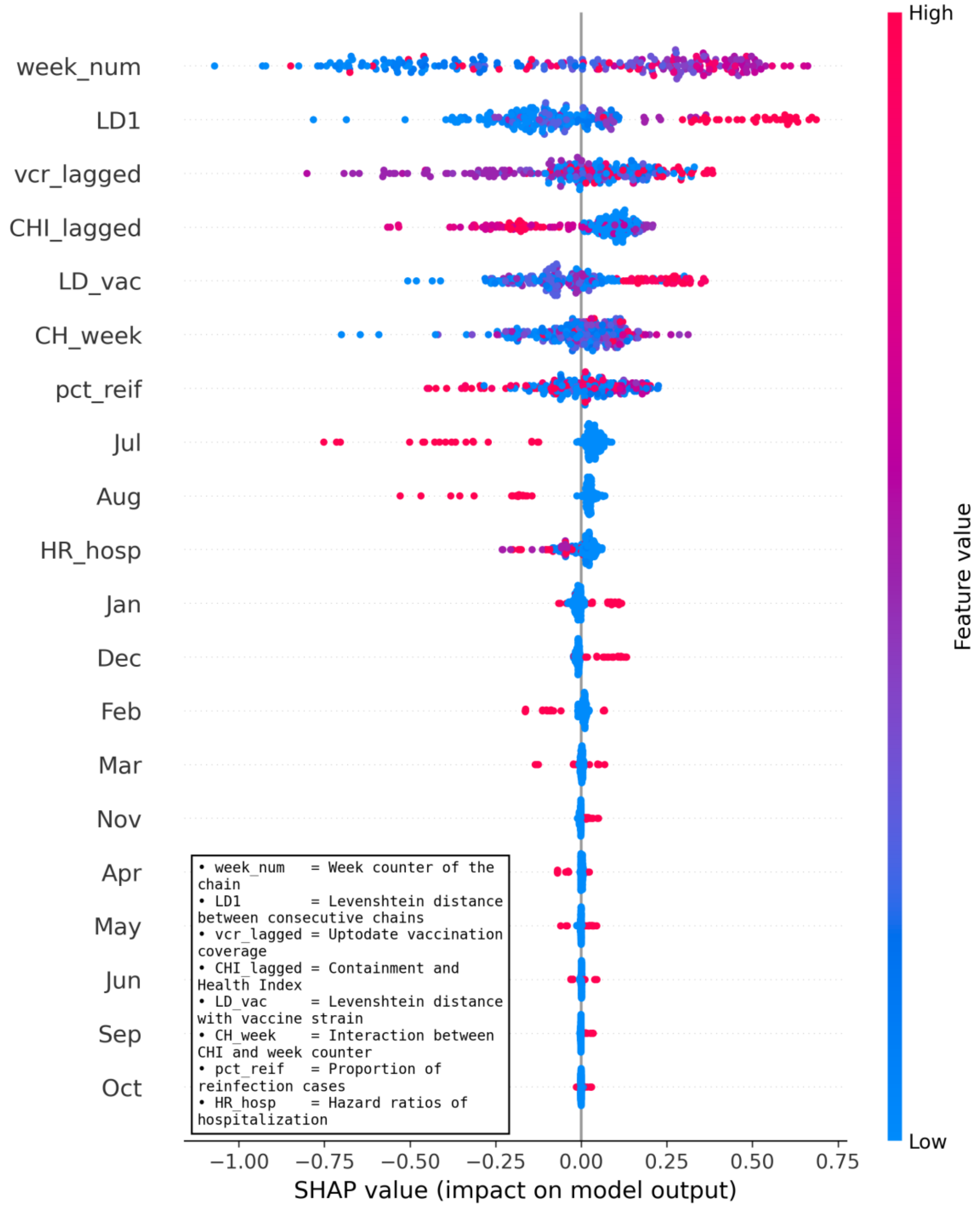


Supplementary Figure S11. SHAP values illustrate variable impact on CatBoost model predictions for weekly hospitalizations in Denmark. Variables are ranked by contribution. High variable values with positive SHAP values indicate positive association, while those with negative SHAP values indicate negative association.

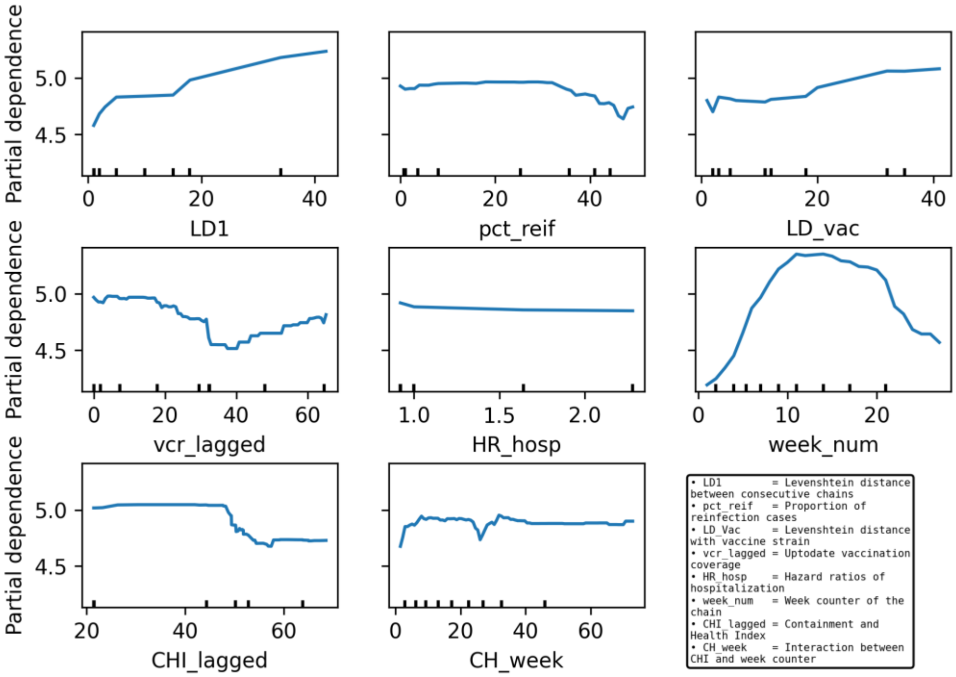


Supplementary Figure S12. Partial dependence plots of selected variables showing the impact of each variable on CatBoost model predictions for weekly hospitalizations in Denmark, while holding all other variables constant.

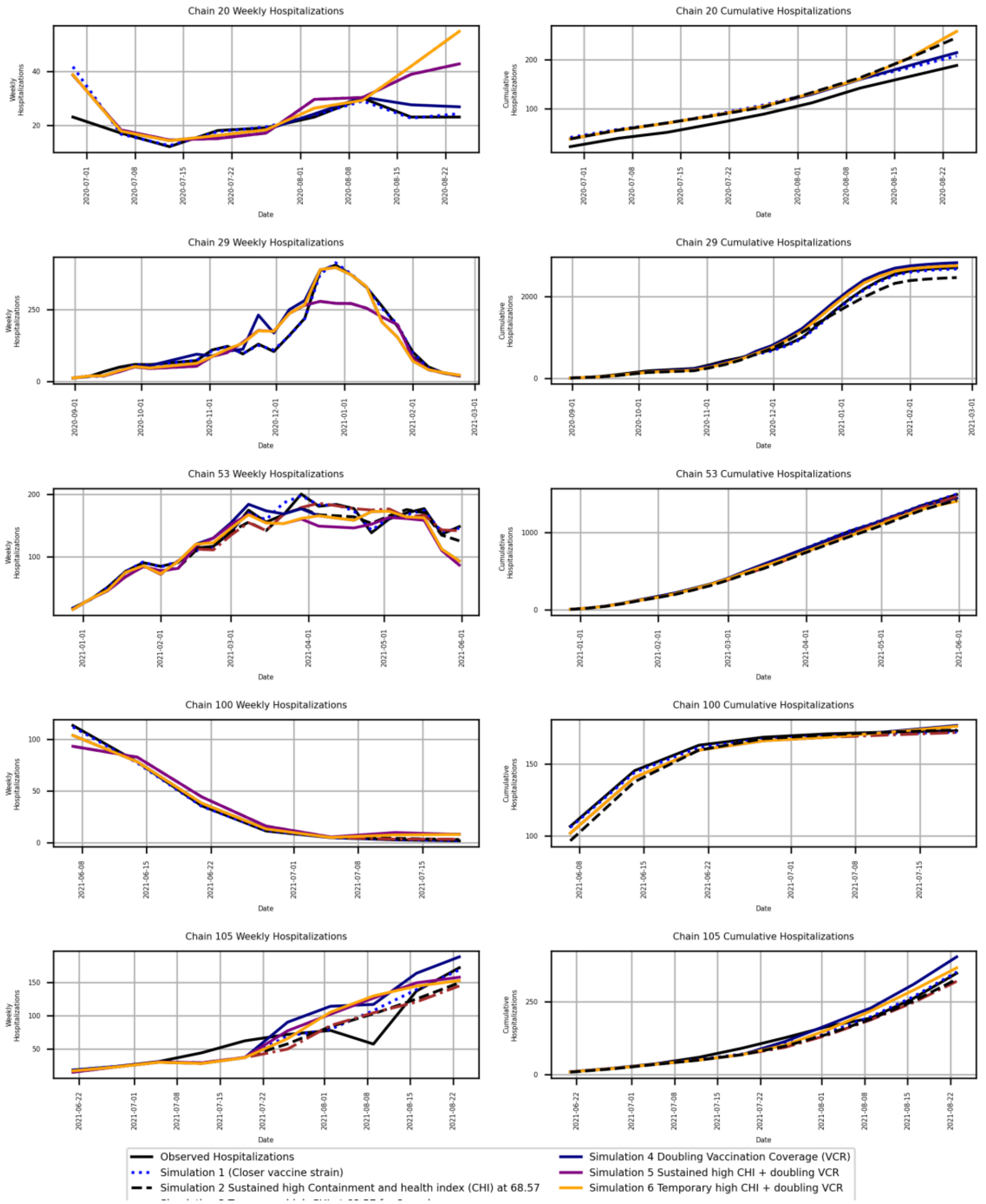


Supplementary Figure S13. Observed hospitalizations and simulation results across various intervention scenarios for weekly and cumulative hospitalizations generated using CatBoost model, from chain 20 to chain 105 in Denmark.

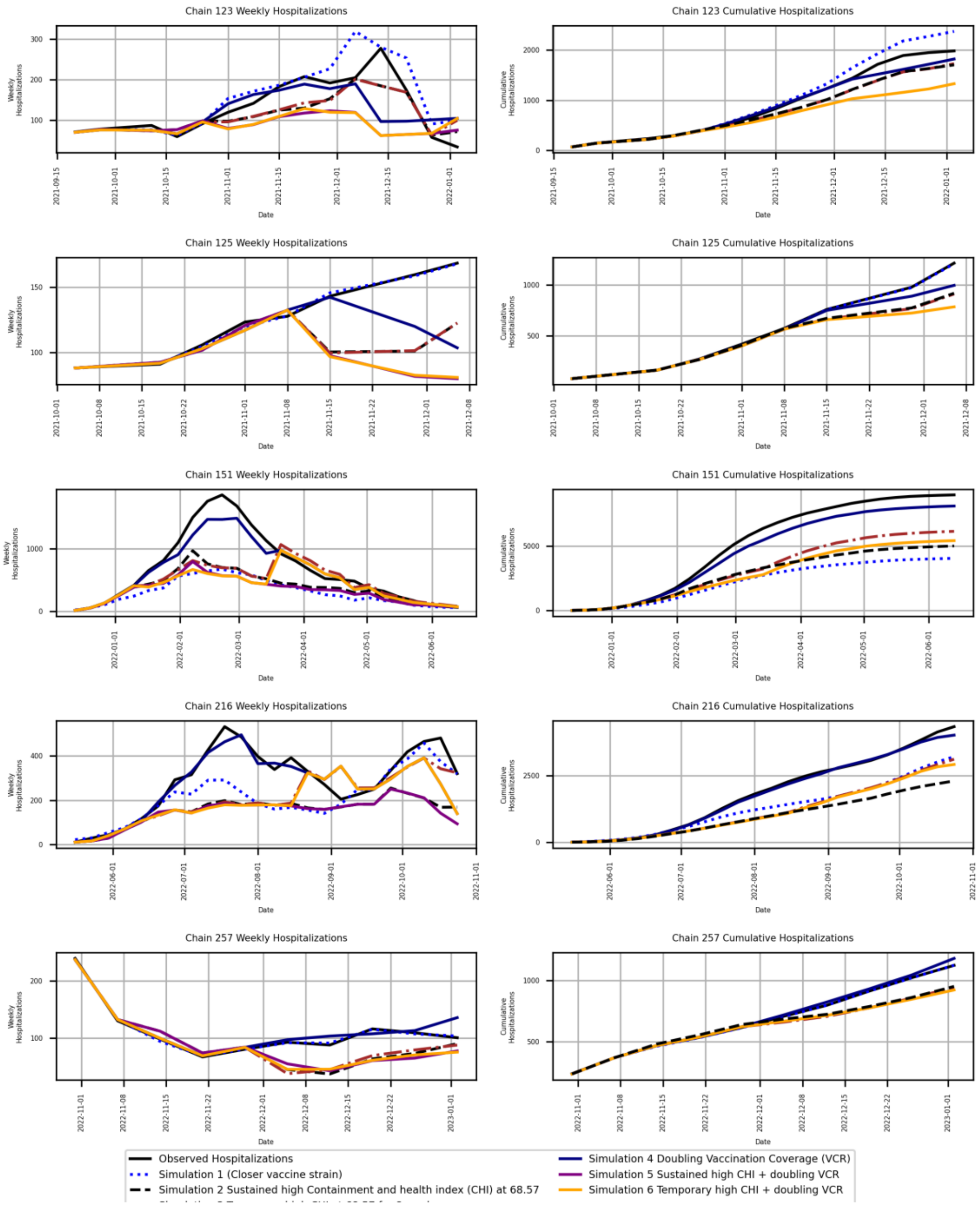


Supplementary Figure S14. Observed hospitalizations and simulation results across various intervention scenarios for weekly and cumulative hospitalizations generated using CatBoost model, from chain 123 to chain 257 in Denmark.

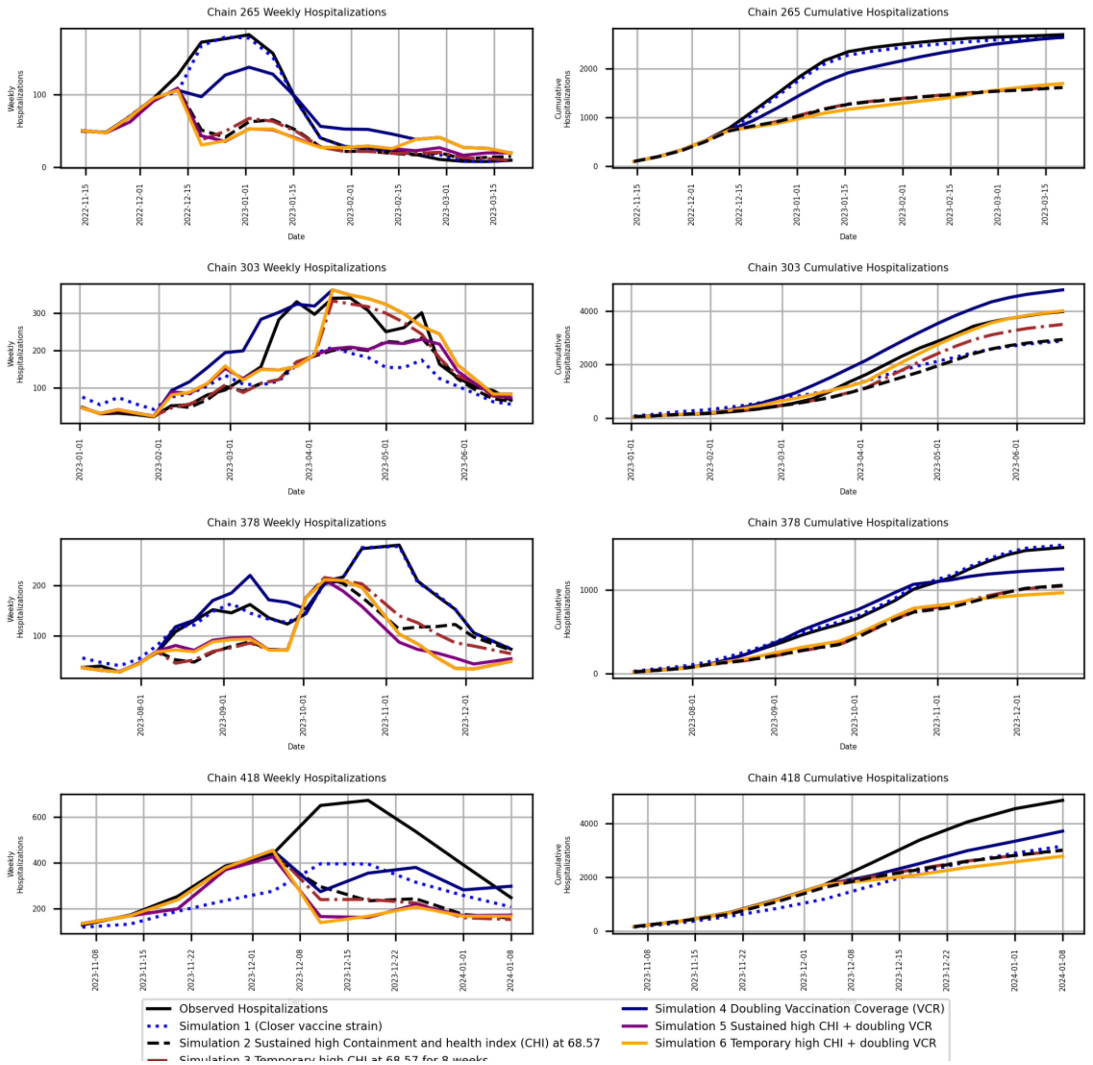


Supplementary Figure S15. Observed hospitalizations and simulation results across various intervention scenarios for weekly and cumulative hospitalizations generated using CatBoost model, from chain 265 to chain 418 in Denmark.
